# Association between social networks and substance use among male tribal adolescents in the North-East Indian state of Tripura

**DOI:** 10.1101/2023.05.07.23289634

**Authors:** Benjamin Debbarma, Ankita Srivastava, Nandita Saikia

## Abstract

**Purpose:** The prevalence of substance use among tribal adolescents in north-east India is higher than that of the rest of India.

**Objective:** The study aimed to investigate the association between social network measures and substance use among male tribal adolescents in the West district of Tripura, North-East India.

**Methods:** We used data on 12-19-year-old tribal adolescents (N=340) from a primary cross-sectional survey in selected schools in the study area. We carried out bivariate and logistic regression analysis to establish the association between substance use and social network.

**Results:** Out of the total sample 340, about 27.65% reported smoking, 26.18% reported using smokeless tobacco, and 30.59% reported drinking alcohol; 35.29% reported using any of these substances. The substance use status of social network members was highly correlated to the substance use status among adolescents. The odds of substance consumption among adolescents increase with having a friend who smokes (OR = 6.152, 95% CI = 1.80–21.09), having friends who instigate to smoke (OR = 5.41, 95% CI = 1.86–15.74), and having friends who say smoking as a sign of masculinity(OR = 5.19, 95% CI = 1.4–18.22). Adolescents were more likely to smoke when their family member uses a substance (OR = 3.39, 95% CI = 1.5– 7.4, *p* = 0.002) and who spent time with friends with the same behaviour (OR = 2.66, 95% CI= 1.5–4.5, *p* ≤ 0.000).

**Conclusions:** Intervention is needed to address adolescents’ substance use habits and members belonging to close social networks.

## Introduction

Substance use refers to the use of any psychoactive substance or drug, including licit and illicit drugs, other than when medically indicated (1,2). The adolescence period has the greatest window of vulnerability for high-risk behaviours, which may continue across the life span (3). Many studies established, early alcohol initiation of substance has been associated with the development of alcohol problems (4,5). Globally, 20 million people initiate substance use at a sensitive period of life (< 14, years of age) which creates a considerable risk for developing substance use disorder at later life (6). Furthermore, substance use poses a threat to an individual’s health, social and economic fabric of families, communities, and nations. Several studies investigated the association of substance use during adolescence with various health and well-being indicators. Substance use during adolescence has been associated with alterations in brain structure, function, and neuro-cognition (7). It is commonly associated with unsafe sexual behaviour, school and social misbehaviour, poor academic performance, and may eventually lead to the continuation of drug use in adulthood (8). It is also the main contributor to the incidence of accidents and injuries during adolescence, representing the leading cause of morbidity and mortality in this age group(9).

Despite the severe threats posed by adolescent’s substance use, substance use among adolescents and youth has remained grossly under-researched in India. As per National Family Health Survey 2015-2016 (NFHS-4), 18.5% boys aged 15-19 years reported tobacco use, and 9% reported alcohol use (10,11). Within India, substance use is higher in North-East regions (2). Studies show that north-eastern states like Arunachal Pradesh, Tripura have the the highest prevalence of alcohol use within India (2,12). A recent study revealed a notable increase in alcohol consumption in Goa, Tamil Nadu, Tripura, and Sikkim where Tripura ranked second highest in the alcohol consumption (13). According to the NFHS-4, 67.8% of men (15-49 age group) in Tripura used any kind of tobacco, while 57.6% of men drink alcohol (10).

Further, previous studies show that the prevalence of alcohol consumption and smoking are high among scheduled tribes (STs) of India, a socio-economically backward population group with the distinct social, cultural, historical, and geographical background (14). Yet detailed study investigating substance use among tribal adolescents are minimum in these states.

Social networks play a crucial role in substance use behaviours (15). Social network refers to networking of individuals or small group in online or offline platforms. Such networks can operate at many levels from the level of families, school, religious organisations or levels of any specific population groups (16). It may refer to the relationship between the members in the network, such as intimate partners, work colleagues, drinking friends, and the level of trust and closeness between them(17). Recent research on alcohol consumption has primarily focused on social networks. Peer influence, specific social group membership, substance behaviour of close relatives, network position are identified as the significant risk factors that influence adolescent’s substance use(18,19). Adolescents are more likely to drink alcohol if their close friends drink alcohol. Encouragement by peer groups, the lure of popularity, and early availability of substances make an adolescent an easy prey (20). Exposure to parental substance use disorder predicts adolescent’s substance use disorder independently of other risk factors (21). However, there is limited literature on the association between substance use and social networks in adolescents from North-east India (2,22).

Our study examined the level of substance use among tribal male adolescents in in West district of Tripura. It also investigated the male tribal adolescent’s social network and its relationship with substances use behaviours in the study area.

## Materials and Methods

### Study design and sampling method

We conducted a primary school-based survey in the selected schools of West district of Tripura between January-April 2019.We purposively selected the West district of Tripura. Out of all schools, we selected four schools having 90% Scheduled Tribe students. Out of total students in each class from IX to XII, 23 students were selected with Simple Random Sampling without replacement. Hence we targeted a total of 92 students from each school. The schools provided the sampling lists for the students from class IX to XII.

We interviewed male tribal adolescents aged 12-19-year-old. Out of the targeted 368 students, 340 students participated in the interview. For each substance, we asked about the use of smoking, smokeless tobacco, and alcohol 30 days prior to the survey. Substance use measures for each question were coded as 1 for use and 0 for no use. Besides, we computed “any substance use” if a student used any smoking, smokeless, or alcohol.

We also collected information on amount of substance use during a single day, age of starting and the duration of use. A 10% random checking of the accuracy of data collection was done by revisiting the study site. Analysis of this data showed no significant error in the data collection.

### Inclusion criteria

a. Age group 12-19 years.
b. Those youth who were willing to participate in the study.
c. Only male Scheduled Tribe youth.

### Exclusion criteria

a. Individuals below 12 years and above 19 years.
b. Incompletely answered questionnaires.
c. Those youth who were suffering from any significant physical/mental instability or unable to provide information.

### Data collection tool

A self-administered cross-sectional anonymous structured questionnaire validated in pilot test were distributed among students. Before filling the questionnaire, students were informed about the study’s anonymity, objective, voluntary nature. The questionnaire was in the English language. We collected and analysed four indicators: smoking tobacco, smokeless tobacco, consuming alcohol, and using any of these three substances among males aged between 12 and 19 years. All of these indicators are dichotomous (Yes/No).

The study has selected four categories or domains of explanatory variables: friends substance use, family member substance use, friendships networks, and usages of social media. These four domains were regarded as social network variables. The details of social networks domain/components are discussed in Appendix Table.

**Friend’s substance use includes i)** friend’s substance use (yes; no); ii) friend’s instigation (yes; no); iii) friend’s substance use perceptions with multiple responses (sign of masculinity; helps in medication; ideal leader/actor does; boost confidence; good for health; reduces boredom).

**Family member substance use** was the information about the parental substance use in the prior to 30 days (yes; no).

**Friendships networks includes i)** groups of friends nominated by the respondents (No, school friends ; any other friends (friends from sports, theatre, dance, sect, and music activities); ii) spent time with friends (very often; frequent and Not at all); iii) reported close friends from (classmate; classmate’s friends; locality); iv) stay together with friends (yes, no, not replied) v) work together with friends (yes, no)

**Usages of social media include** use of social networking sites (SNS) such as WhatsApp and Facebook.

We have controlled the role of other socio-economic factors such as father’s education (illiterate, primary, secondary and higher), mother’s education (illiterate, primary, secondary and higher), family income (do not know, 800-2000, 3000-6000), and type of ration card owned by the family. A ration card is the official document issued by the respective state governments, enabling eligible households to buy food grains at subsidized rates under the National Food Security Act (23). The document serves as a common form of identification for many individuals. Different types of ration cards in India includes Priority household (PHH) ration card, issued to those households which meet the eligibility criteria set by the state governments. These households entitled to 5 kilograms of food grains per member every month. Antyodaya Anna Yojana (AAY) ration card is issued to those households which fall under the ‘poorest of poor’ category. AAY card holders are entitled to 35 kilograms of food grain every month. Below Poverty line (BPL) ration card was for those households living under the poverty line. Above Poverty line (APL) ration cards were issued to households living above the poverty line (as estimated by the <u>Planning Commission).</u> These households received 15 kilogram of food grain (based on availability) (24).

## Statistical analysis

Statistical analysis was carried out using STATA version 13.1. Categorical variables were presented as frequencies and percentage. The response variables—types of substances use were dichotomous variables categorized as yes and no. To measure the association between social networks variables and substance use, we used binary logistic regression. Odds ratios and confidence intervals were computed. Statistical significance was set at P < 0.05 for the study.

### Ethics statement

The present study is a primary school-based survey in the selected schools of West district of Tripura between January-April 2019, including male tribal adolescents aged 12-19-year-old. The survey protocol was approved by the Ethical Review Committees, Institutional Ethics Review Board of the Jawaharlal Nehru University (IERB-JNU). Informed assent and parental consent were obtained for male tribal adolescents aged 12–19 years, and written informed consent was obtained from the school authority.

## Results

### Descriptive and bivariate characteristics

#### Age of initiation and prevalence of substances use

Figure 1 shows the prevalence of smoking, smokeless tobacco, alcohol, and any substance use during one month prior to the survey. Out of the total sample 340, about 27.65% reported smoking, 26.18% reported using smokeless tobacco, and 30.59% reported drinking alcohol; about 35.29% reported using any substances.

**Figure 1.**
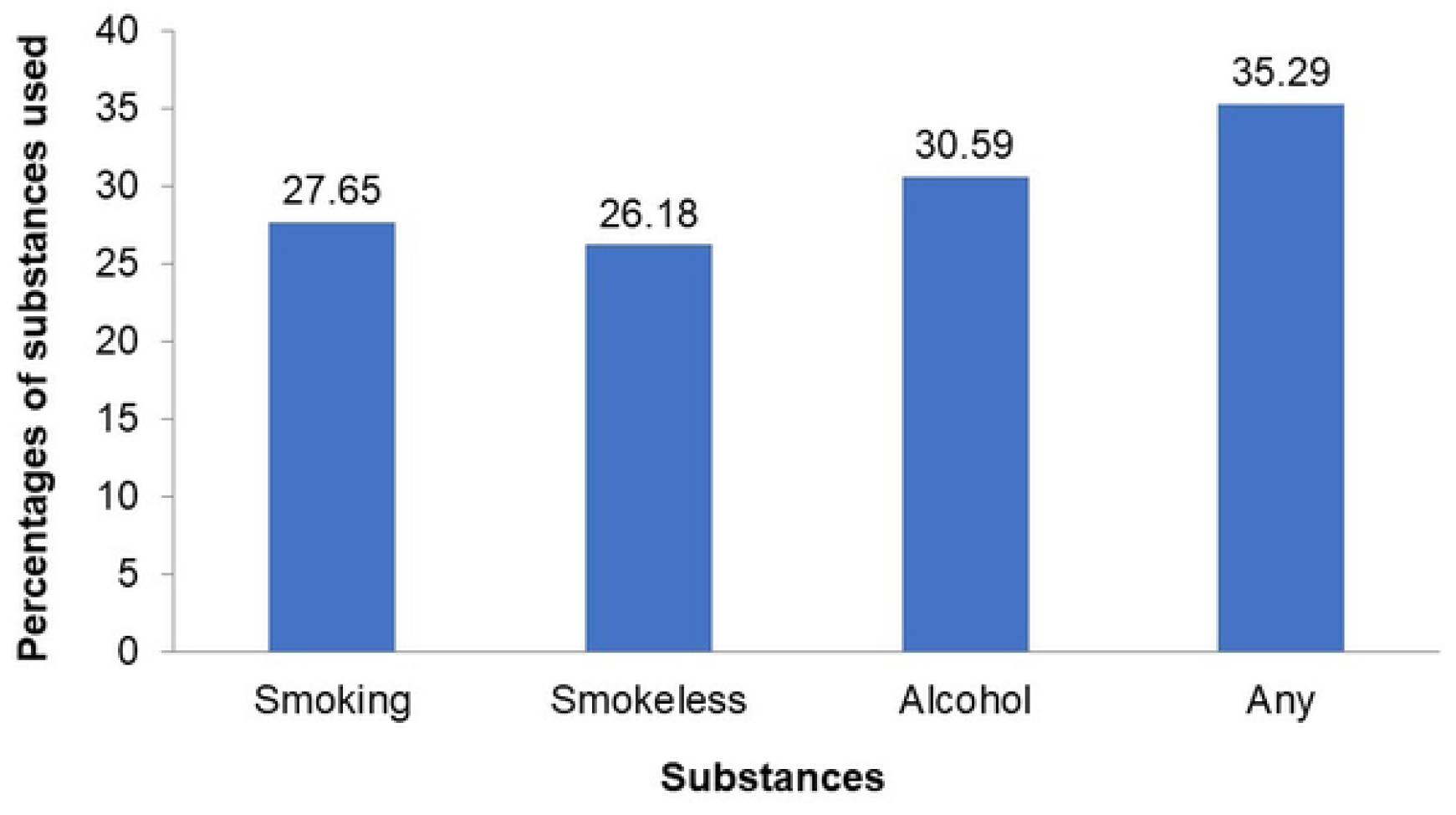
Prevalence of substances used among adolescents aged 12-19 years, West District, Tripura, 2019. Source: Primary field survey, January-April 2019

Figure 2 shows the age at initiation of substance use among the students. Substance use initiation has been measured by asking about the age of initiation for smoking, smokeless tobacco, and alcohol consumption. Out the total respondents, the most common age of substance initiation was 14-18 years, the lowest proportion of substance initiation was reported in 7-14 years. While less than 22% students initiated alcohol or smoking in age 7-14, about 91.18 % of them initiated smokeless tobacco at the age of 7-14 years.

**Figure 2.**
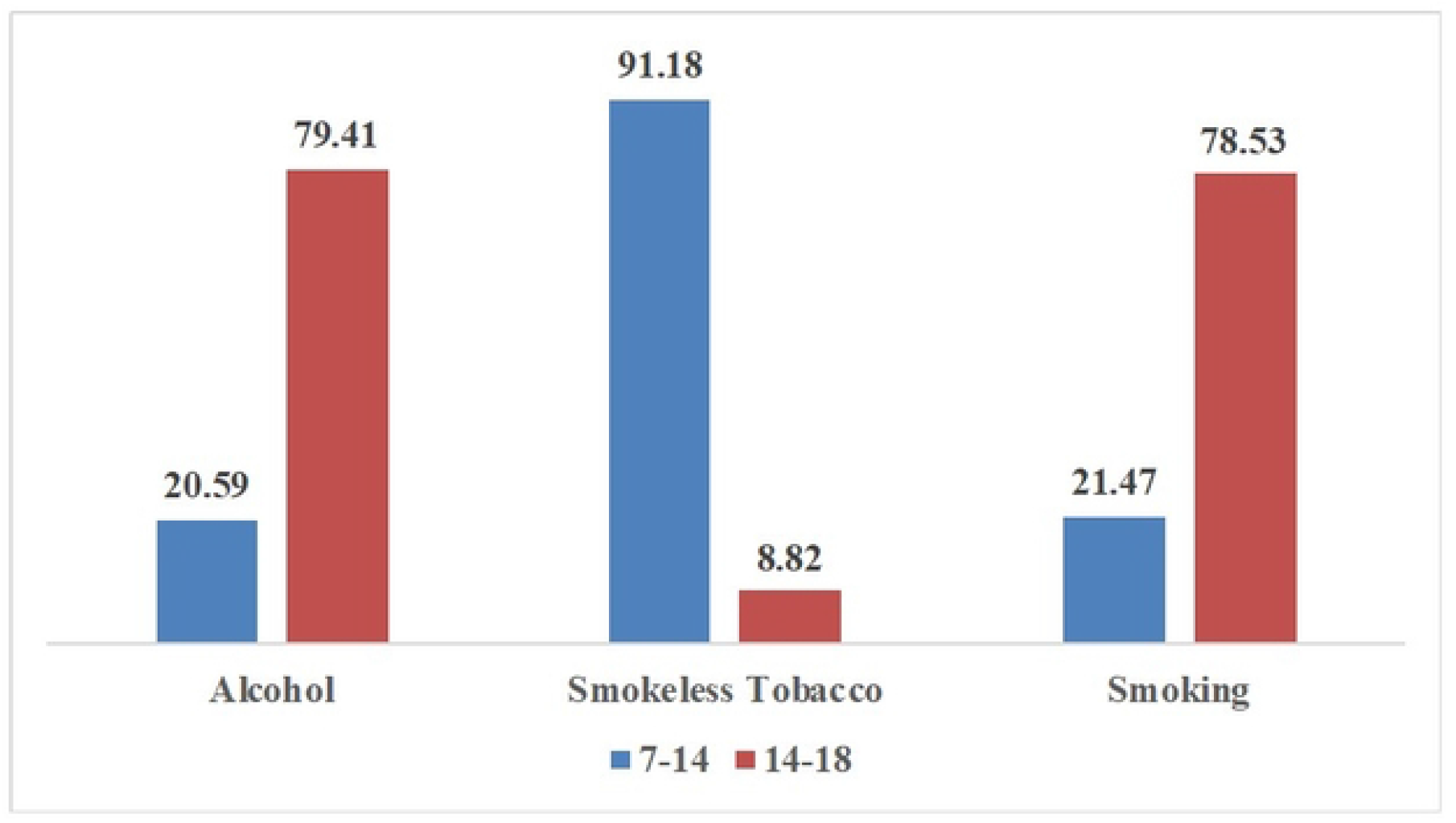
Age at first use of substances among male tribal adolescents in West district of Tripura, 2019. Source: Primary field survey, January-April 2019

#### Association between Social network and adolescent’s substances consumption

Table 1 presents the descriptive characteristics of the studied sample. About 73% of the respondent’s fathers were secondary or above educated; whereas 53.53% of the respondents’ mothers attained at least secondary level of education. About 52.06% of the respondent’s household income was above Rs 7000 per month, and about 23.82% of the respondent’s household lived below the poverty level.

**Table 1.**
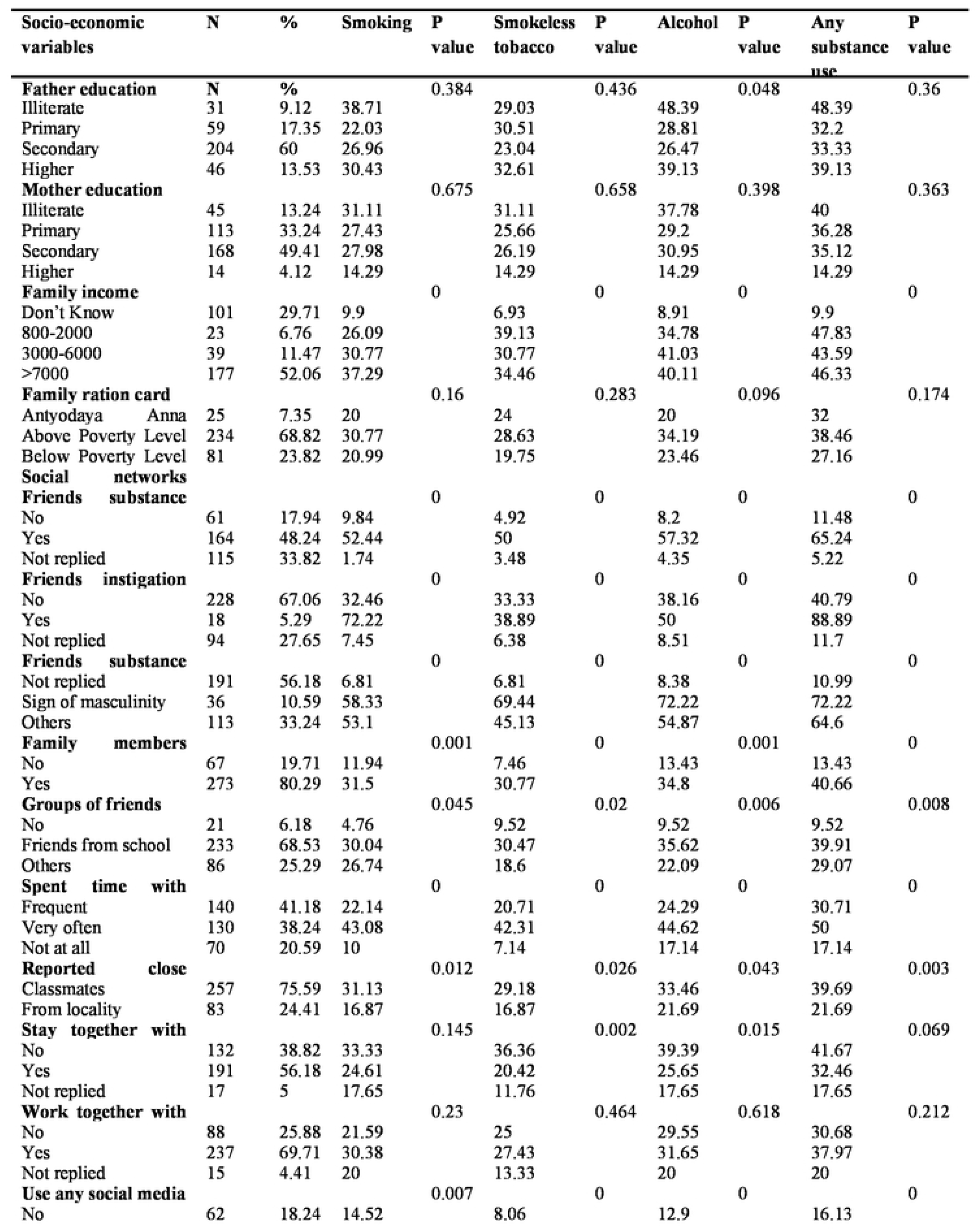
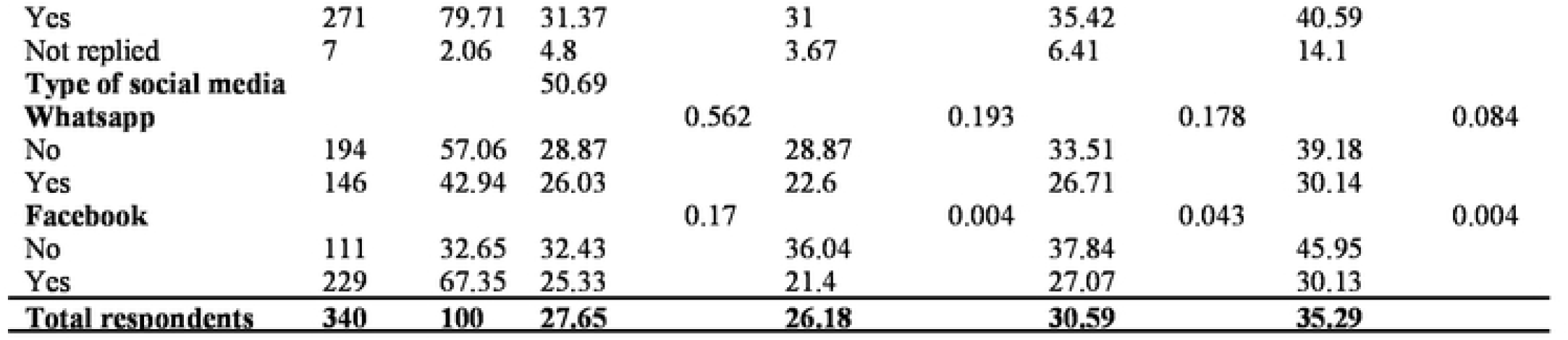
Bivariate analysis of substances consumption with descriptive characteristics, among young male youths aged 12-19 years (n=340), West district Tripura, 2019

About 48.24% of the respondent’s friend consumed substances whereas 80.29% of the family members of the respondents’ consumed substances. Nearly 80% of the students spent time with their friends very often or frequently; majority of them have friends from the class (75.59%). About 79% of the respondents used social media; 42.94% used social media platform called “WhatsApp” whereas 67.35% used “Facebook”.

Table 1 also describes the association between substance use and socio-economic and demographic characteristics. Substance use generally increases with the increasing social network. Any substance use increased significantly when friends used substance (65.24 vs 11.48, *p* ≤ 0.000) and when friends instigated to do so (88.89 *vs*. 40.79, *p* = 0.000). Substance use also increases when friends perceive it as a sign of masculinity (72.22 vs. 10.99, p≤0.000). It was also significantly associated with family member’s substance use (40.66) and time spent with school friends with same behavior (50.00), *p* ≤ 0.000). Substance use is higher among social media users compared to non-users (40.59 vs. 16.13, p=0.000).

Specifically, adolescents’ smoking (or smokeless tobacco status or alcohol consumption) is significantly associated friend’s substance use (52.44% *vs*. 9.84%, *p* ≤ 0.000). Besides it increased with instigation by friends for all three categories, say, in case of smoking, it is 72.22% *vs*. 32.46%, *p* ≤ 0.000; in case of smokeless tobacco, it is 38.89% *vs*. 6.38%, *p* ≤ 0.000) and in case of alcohol consumption, it is 0.00% *vs*. 8.51%, *p* ≤ 0.000. Friend’s perception to use the substance as a sign of masculinity also played a significant role in the consumption of alcohol, in smokeless tobacco and in smoking. Substance use of all types increased if the family member is a substance user. Adolescents who spent time with friends consume more substances than the rest; smoking (43.08% *vs*. 10.00%, *p* ≤ 0.000), smokeless tobacco (42.31% *vs*. 7.14%, *p*=0.000), and alcohol consumption (44.62% vs. 17.14%). Substance use was also found significantly high when group of friends are from school. Furthermore, adolescent’s using any social media has higher prevalence of substance consumption. (Table 1). Other study variables including mother’s and father’s education and family ration cards do not significantly affect adolescents’ substances consumption behavior.

The binary logistic regression analysis results for each type of substance use (smoking, smokeless tobacco, alcohol or any substances use) are presented in Table 2. The results of logistics regression analysis demonstrate that social network characteristics of adolescent’s’ are significantly associated with tobacco and alcohol consumption.

**Table 2.** Binary logistic regression analysis^1^ of substances consumption with social networks characteristics in the West district of Tripura, India, 2019

| Social networks | Smoking<br>OR (95% CI) | p- | Smokeless tobacco<br>OR (95% CI) | p- | Alcohol<br>OR (95% CI) | p- | Any substance<br>OR (95% CI) | p- |
| --- | --- | --- | --- | --- | --- | --- | --- | --- |
| **Friends' substance use** |  |  |  |  |  |  |  |  |
| No® | 1 |  | 1 |  | 1 |  | 1 |  |
| Yes | 6.15(1.8-20.2) | 0.009 | 0.61(0.17-2.18) | 0.39 | 6.16(1.95-19.3) | 0.001 | 8.62(2.74-27.4) | 0.000 |
| Not replied | 0.2(0.03-1.5) | 0.09 | 0.46(0.08-2.4) | 0.39 | 0.34(0.07-1.6) | 0.17 | 0.32(0.07-1.5) | 0.12 |
| **Friends' instigation of substance use** |  |  |  |  |  |  |  |  |
| No® | 1 |  | 1 |  | 1 |  | 1 |  |
| Yes | 5.41(1.86-15.4) | 0.003 | 0.6(0.16-2.1) | 0.44 | 0.98(0.28-3.5) | 0.96 | 5.11(0.8-30.8) | 0.08 |
| Not replied | 0.17(0.07-0.4) | 0 | 1.45(0.4-5.2) | 0.57 | 1.52(0.47-4.9) | 0.49 | 2.25(0.75-6.8) | 0.15 |
| **Friends' substance use perception** |  |  |  |  |  |  |  |  |
| Sign of masculinity® | 1 |  | 1 |  | 1 |  | 1 |  |
| Not replied | 5.19(1.46-18.4) | 0.01 | 1.22(0.57-2.67) | 0.62 | 15.57(4.21-58.1) | 0.000 | 6.36(1.7-23.8) | 0.004 |
| Others | 4.79(1.78-12.8) | 0.003 | 3.87(1.31-10.9) | 0.01 | 6.46(2.62-16.1) | 0.000 | 5.3(2.16-13) | 0 |
| **Family members substance use** |  |  |  |  |  |  |  |  |
| No® | 1 |  | 1 |  | 1 |  | 1 |  |
| Yes | 3.39(1.55-7.4) | 0.002 | 4.8(1.28-17.4) | 0.02 | 1.91(0.67-5.3) | 0.23 | 2.51(0.9-6.6) | 0.08 |
| **Groups of friends** |  |  |  |  |  |  |  |  |
| No® | 1 |  | 1 |  | 1 |  | 1 |  |
| Friends from school | 8.59(1.13-65.4) | 0.04 | 1.36(0.13-14.1) | 0.8 | 7.43(0.78-70.1) | 0.08 | 6.82(0.73-63.4) | 0.09 |
| Others | 7.3(0.93-56.1) | 0.06 | 1.58(0.17-14.4) | 0.69 | 4.18(0.48-36.1) | 0.2 | 4.65(0.54-39.4) | 0.16 |
| **Spent time with friends** |  |  |  |  |  |  |  |  |
| Frequent® | 1 |  | 1 |  | 1 |  | 1 |  |
| Very often | 2.66(1.57-4.5) | 0.000 | 2.04(0.89-4.6) | 0.09 | 1.84(0.83-4.1) | 0.13 | 1.36(0.59-3.1) | 0.47 |
| Not at all | 0.39(0.16-0.9) | 0.04 | 0.17(0.04-0.7) | 0.02 | 0.59(0.2-1.74) | 0.34 | 0.27(0.09-0.7) | 0.02 |
| **Reported close friends from** |  |  |  |  |  |  |  |  |
| Classmates® | 1 |  | 1 |  | 1 |  | 1 |  |
| From locality | 0.45(0.24-0.8) | 0.01 | 0.62(0.19-2.0) | 0.44 | 1.43(0.48-4.3) | 0.53 | 0.97(0.32-3.0) | 0.96 |
| **Stay together with friends** |  |  |  |  |  |  |  |  |
| No® | 1 |  | 1 |  | 1 |  | 1 |  |
| Yes | 0.65(0.4-1.0) | 0.09 | 0.45(0.27-0.74) | 0 | 0.53(0.33-0.84) | 0.01 | 0.67(0.43-1.0) | 0.09 |
| Not replied | 0.43(0.12-1.5) | 0.2 | 0.23(0.05-1.0) | 0.06 | 0.33(0.09-1.2) | 0.09 | 0.71(0.51-1.0) | 0.06 |
| **Work together with friends** |  |  |  |  |  |  |  |  |
| No® | 1 |  | 1 |  | 1 |  | 1 |  |
| Yes | 1.59(0.89-2.8) | 0.12 | 1.13(0.65-1.9) | 0.66 | 1.1(0.65-1.88) | 0.72 | 1.38(0.82-2.3) | 0.23 |
| Not replied | 0.91(0.23-3.7) | 0.89 | 0.46(0.1-2.1) | 0.33 | 0.6(0.16-2.29) | 0.45 | 0.57(0.15-2.1) | 0.41 |
| **Use any social media** |  |  |  |  |  |  |  |  |
| No® | 1 |  | 1 |  | 1 |  | 1 |  |
| Yes | 2.69(1.27-5.7) | 0.01 | 5.12(1.98-13.1) | 0 | 3.7(1.69-8.1) | 0 | 3.55(1.73-7.3) | 0 |
| Not replied | 0.17(0.08-0.3) | 0 | 0.09(0.04-0.2) | 0 | 0.15(0.07-0.3) | 0 | 0.19(0.1-0.3) | 0 |
| **Types of social media** |  |  |  |  |  |  |  |  |
| **WhatsApp** |  |  |  |  |  |  |  |  |
| No® | 1 |  | 1 |  | 1 |  | 1 |  |
| Yes | 1.27(0.57-2.8) | 0.56 | 1.34(0.56-3.2) | 0.51 | 0.92(0.41-2.1) | 0.83 | 0.78(0.34-1.8) | 0.55 |
| **Facebook** |  |  |  |  |  |  |  |  |
| No® | 1 |  | 1 |  | 1 |  | 1 |  |
| Yes | 1.29(0.55-3) | 0.56 | 1.11(0.46-2.7) | 0.81 | 1.12(0.48-2.6) | 0.79 | 0.56(0.23-1.3) | 0.21 |

<sup>1</sup> All models are controlled for the other socio-economic variables presented in table 2.

The result shows that smoking has six times higher odds if the user has a friend who smokes (OR = 6.152, *p* ≤ 0.000). Odds of smoking were five times higher if friends instigate to smokes (OR = 5.41*p* = 0.002). One highlight of the findings is that friend’s perception of smoking as a sign of masculinity has five times higher odds (OR = 5.19*p* = 0.000). Moreover, adolescents were more likely to smoke if family member smoked (OR = 3.39*p* = 0.002), and spent time with friends of the same behaviour (OR = 2.66*p* ≤ 0.000) (see Table 2).

Smokeless tobacco consumption among adolescent men had lower risk, if his friends consume (OR= 0.61, *p* ≤ 0.000). Friends’ instigation had precisely half the risk of smokeless tobacco consumption (OR= 0.59, p=0.000). However, friends’ perception of smokeless tobacco has 1.2 times higher odds (OR= 1.21, p = 0.000), and adolescents with addicted family members had about 5 times higher odds of smokeless tobacco (OR= 4.80, *p* = 0.000). Social media exposed user had 5 times higher risk of having smokeless tobacco (OR= 5.12, *p* = 0.000). The odds of smokeless consumption with groups of friends, time spent, close friends, staying or working together with friends were not statistically significant.

The results for alcohol consumption showed friends’ substance use (OR=6.16, *p* = 0.000) and friend’s perception (OR=6.45, *p* = 0.000) had six times higher odds of alcohol consumption, while friend’s instigation (OR= 0.97, *p* ≤ 0.000), any social media exposure (OR=0.14, *p* ≤ 0.000) have lower odds of alcohol consumption among adolescents; and with family members uses adolescent had 1.91 times higher odds (OR= 1.91, *p* ≤ 0.000). These effects were statistically significant. Male respondents with friends had five times higher odds of consuming any substance use (OR=5.29, p=0.000). Respondents having any social media exposure and social media account were statistically significantly associated with any kind of substance use.

## Discussion and conclusion

Substance abuse is a major public health concern in the world (25).The effects of drug usage are often multifaceted. This habit affects health, education, and occupational career and incurs a huge financial and social burden that affects health, education, and occupational career and incurs a huge financial and social burden to society (8). Early substance use is typically associated with poor prognosis and a lifelong history of deception and reckless behaviour(26). It is therefore crucial to identify the risk factors of substance consumption among adolescents. The present study investigated the connection between social networks and substance use among male tribal secondary school students aged 12-19 in Tripura’s West District.

The high prevalence of smoking, smokeless tobacco, alcohol and any of these substances among male tribal adolescents in Tripura is alarming. While 18.5% boys aged 15-19 years reported tobacco use, and 9% reported alcohol use at national level, the prevalence of any substance use among study population was as high as 35.29%. Previous studies conducted in other parts of the world have unequivocally established that initiation of drug use usually occurs between the ages of 12 and 24 years (27). Our study findings reiterate earlier findings among male tribal youths in Tripura.

Our results show that friend’s substance use status is an influential factor in adolescent substance use behaviour as it’s effect remained significant even after controlling the role of other factors. Similar findings have been observed in other studies (28). As they mature, friends become increasingly crucial to teenagers (29). As adolescents become more autonomous, the peer group’s influence becomes more important (30). A significant number of studies have reported that peers plays major role in the growth of adolescents’ risk behaviour.The study also supports the finding that adolescents who had friends using drugs had an effect on them, encouraging or discouraging risky behavior(31). Among teenagers, the pressure to gain acceptance among friends is normal. Understanding peer influencewould encourage practitioner to design effective interventions to prevent youth from developing risky behaviour (32).

Besides social influence, peer or friends as determinants of substances use, modern information and communication technologies such as, SNS is highly popular among adolescents and played an important role (33). With SNS usage, adolescent were exposed to unhealthy substances and advertisement in digital media (34). In their formative years, social media addiction can have a deleterious impact on the user’s physical health, psychological health, and behavioural issues. Researchers have found that 25-37 percent of older adolescents post information about their alcohol intake in social media(35).

The online exchanges with friends might mediate peer influence processes (regarding adolescent cigarette and alcohol use) by conveying information about peers’ risk-taking behaviour (36). Extensive research has been undertaken to measure and understand the impact of social media portrayals of tobacco use on young people’s tobacco related attitudes and behaviours (37). Especially within the context of adolescents, where exposure to social media are being a central to social order in many ways (38).The peer influence has been established as a leading correlation of adolscent drug use be hvior (32).Previous research indicates the association between substance usage by friends who consume alcohol and use of social media, which may suggest that online interactions with friends could mediate peer control mechanisms (in terms of adolescents’ cigarette and alcohol use) by sharing knowledge about the risk-taking behaviour of peers (36).

Family has a significant role in adolescents’ psychological well-being and health risk behaviours (39). A wide range of family factors, including family structure, family process, parenting styles, and family members’ smoking habits be associated with adolescent smoking and drinking (40). This study shows that adolescents whose family members consume substances were more likely to influence them. Studies have found out that parental education and income were associated with higher rates of substances use. Higher parental education and income are correlated with higher rates of binge drinking in young adults (41).

There are a few limitations of this study. First, the study’s population was not representative of the entire adolescent population in Tripura and West district. Participants were drawn from a tribal dominated district with rural and semi-urban families of low socio-economic status. Second, the study’s cross-sectional nature limits our ability to make causal inferences on the assessed variables. Third, the analyses conducted were based on self-reported data, making response bias a possibility.

### Policy implications

Parents, school authorities and healthcare professionals in the study area should be cautioned about the rise of such activities. Youth drinking behaviors should be closely monitored via school, religious institutions and other non-governmental organization. In this study, the substances use status of social, family members, friends or peers and social media were all correlated in developing smoking and drinking habits among adolescents. Friends of smokers are far more likely to be substance user than friends of non-smokers/non-drinkers. Familial influences are also seen as essential factors in the development of adolescents. Children look up to their parents as role models. Therefore, intervention is needed to address adolescents’ and substance use habits and close social networks members.

## Data Availability

All relevant data are within the manuscript and its Supporting Information files.

## Data Availability

All relevant data are within the manuscript and its Supporting Information files.

## Appendix Table Definition of the variable’s substance use, social media and peer groups

**Appendix Table Definition of the variable's substance use, social media and peer groups**
| **Variables** | **Questions** |
| --- | --- |
| **Friends' substance use** | Do you have friends who consume substances? (Yes/No) |
|  | Do any of your friends instigate you to consume substances? |
|  | How does your friends circle perceive about substances consumption? |
| **Family member substance use** | Do you have a family member who uses substances? (Yes/No) |
| **Friendships networks** | How often do you spend time with each other's? (1=Frequent, 2= Very often and 3= Not at all) |
|  | Do you have a groups of friends with whom you roam around, spent time or do common activities? (No, Yes and school friends; yes and any other friends/friends from sports, theatre, dance, sect, and music activities) |
|  | Who are your close friends? (1=classmate, 2= classmate friends and 3= from locality) |
|  | Do you stay with any of them together? (Yes/No) |
|  | Do you work together? (Yes/No) |
| **Usages of social media** | Do you use any social media? (Yes/No) |
|  | Which are the social media you use? (Yes/No) |
|  | Whats-app (Yes/No) |
|  | Face-book (Yes/No) |

**Source: Authors elaboration from field survey data, 2019.**

## Footnotes

1 All models are controlled for the other socio-economic variables presented in table 2.

## Notes

### Competing Interest Statement

The authors have declared no competing interest.

### Funding Statement

The author(s) received no specific funding for this work.

### Author Declarations

The survey protocol was approved by the Ethical Review Committees, Institutional Ethics Review Board of the Jawaharlal Nehru University (IERB-JNU). Informed assent and parental consent were obtained for male tribal adolescents aged 12-19 years, and written informed consent was obtained from the school authority.

